# Real-World COVID-19 Vaccine Protection Rates against Infection in the Delta and Omicron Eras

**DOI:** 10.1101/2022.09.01.22279492

**Authors:** Yuru Zhu, Jia Gu, Yumou Qiu, Song Xi Chen

**Author notes:** corresponding authors **Correspondence** Correspondence and requests for materials should be addressed to Song Xi Chen and Yumou Qiu.

## Abstract

The real-world protection rates of vaccination (VPRs) against the SARS-Cov-2 infection are critical in formulating future vaccination strategies against the virus. Based on a varying co-efficient stochastic epidemic model, we obtain seven countries’ real-world VPRs using daily epidemiological and vaccination data, and find that the VPRs improved with more vaccine doses. The average VPR of the full vaccination was 82% (SE: 4%) and 61% (SE: 3%) in the pre-Delta and Delta-dominated periods, respectively. The Omicron variant reduced the average VPR of the full vaccination to 39% (SE: 2%). However, the booster dose restored the VPR to 63% (SE: 1%) which was significantly above the 50% threshold in the Omicron-dominated period. Scenario analyses show that the existing vaccination strategies have significantly delayed and reduced the timing and the magnitude of the infection peaks, respectively, and doubling the existing booster coverage would lead to 29% fewer confirmed cases and 17% fewer deaths in the seven countries compared to the outcomes at the existing booster taking rates. These call for higher full vaccine and booster coverage for all countries.

---

The SARS-Cov-2 has been circulating globally with a sequence of emerging variants since the start of the pandemic. Particularly, the Delta and Omicron variants have contributed to surges in the infected cases across the globe due to their high transmissibility^1, 2^. To prevent the spread of COVID-19, vaccines have been rolled out since later 2020, while the booster shots were started since June 2021. Clinical trials or observational studies have been made to evaluate the effects of a vaccine or an arrangement of mixed vaccines^3–9^. It is found that the vaccine efficacies of the two-dose vaccination against the original SARS-Cov-2 strain ranged from 50.7% to 95%^3, 4, 10^, but waned against Delta and Omicron variants. The vaccine effectiveness ranged from 82.8% to 94.5% against Delta and 48.9%-75.1% against Omicron for two doses of Pfizer, AstraZeneca or Moderna vaccines^8^. The vaccine efficacy is defined as one minus the relative risk in the randomized controlled clinical trials^11^, and the vaccine effectiveness is valued in observational studies, which is one minus the hazard ratio in cohort studies^6^ and one minus the odds ratio in case-control studies^5^. Observational studies were more common in the Omicron era.

The booster dose had been shown to increase protection against infection. For homologous or heterologous booster doses of Pfizer, AstraZeneca and Moderna, the effectiveness was 82.3%-97.0% against Delta and 55.6%-73.9% against Omicron, with higher effectiveness using Moderna as the heterologous booster dose^8^. The vaccine effectiveness was 51.0% for three doses of Sinovac against Omicron^12^, which increased to 63.6% by Sinovac as primary with one Pifzer booster ^13^. See Table S1 in the supplementary for the detailed vaccine efficacy and effectiveness discovered by the existing clinical and observational studies. However, the real-world performance of vaccines at the country’s population level which we call the vaccine protection rate is largely unknown.

Different from vaccine efficacy and effectiveness, the real-world vaccine protection rate (VPR) is defined as one minus the percentage reduction in the infection rate of the vaccinated relative to the unvaccinated population of a country. The VPR measures the combined effectiveness of vaccines administrated in a country at a particular age distribution and nonpharmaceutical intervention measures against COVID-19. The impacts of these factors are not necessarily evaluated in the homogeneous clinical trials, cohort studies or case-control studies. Indeed, the conventional vaccine efficacies are pegged to a specific vaccine or a mix of vaccines in the clinical trials after excluding certain part of the population, which may not conform to the population characteristics of the country. Therefore, the available vaccine efficacy or effectiveness does not necessarily reflect the vaccine immunity level of the whole population against different variants of the SAR-COV-2 virus. Hence, it is useful to obtain the real-world VPRs of a country.

Using the daily epidemiological and vaccination data, which include the cumulative numbers of confirmed cases, deaths, recoveries, and people having received the partial, full and booster vaccination, we construct a varying coefficient stochastic epidemic model with eleven compartments (flow diagram in Figure 4) and develop an estimation procedure for the real-world VPRs as well as the key parameters quantifying the dynamic infection, death and recovery rates, which comprehensively reflects the COVID-19 dynamics and nonpharmaceutical interventions (NPI). Compared with existing studies on the effect of vaccination^14, 15^, we do not assume permanent and full immunity of the vaccines and previous infection while incorporating the stochastic natures of the epidemics with time-varying infection rate due to varying levels of NPI and self protective measures, and allow asymptomatic infection, infection before clinic confirmation, vaccine breakthrough, reinfection and different levels of immunity induced by different vaccine doses. The non-parametric time-varying infection rate in our model is better suited for the COVID-19 pandemic as both the virus transmission rate and the NPI measures change over time.

We considered seven countries that are representative for different types of vaccines with sufficient number of confirmed cases (more than 10% of the total population) after vaccination. Specifically, the results of the US may be used to show the effect of mRNA vaccines (Pfizer and Moderna); the three European countries, UK, Italy and Germany, mainly used a mixture of non-replicating viral vector vaccines (AstraZeneca) and mRNA vaccines; the two south American countries, Brazil and Peru, utilized the inactivated vaccines (Sinopharm and Sinovac), AstraZeneca and Pfizer; Turkey used the inactivated vaccines at the beginning and then started Pfizer. In this paper, the full dose means one dose for Janssen and two doses for the other brands to complete the primary vaccination. Those who have not completed the full vaccination are called partially vaccinated. The booster shot means one dose after full vaccination. The coverage rates of the partial, full and booster vaccines in population are reported in Figure 3 (a), which shows 62.3%–79% of the population in the seven countries have taken the full shots on March 15 2022, but the coverage rates of booster shots were much lower, ranging between 29.1% in the US and 63.4% in Italy.

## Results

For each country, we estimate the vaccine protection rates in six consecutive non-overlapping post-vaccine periods: the pre-Delta, Intervening I, Delta-dominated, pre-Omicron, Intervening II and Omicron-dominated periods. Details of these periods are provided in the method section.

### Vaccine protection rates

The real-world VPRs for the partial, full and booster vaccination in the seven countries in the six post-vaccine periods are reported in Figure 1 with detailed numerical values in Table S3. It shows that before the booster vaccination, both the partial and full vaccination were largely protective against the COVID infection in the pre-Delta period with the VPRs in the seven countries ranging 48%-64% and 68%-95% for the partial and full vaccination, respectively. However, the Delta variant had caused waning VPRs of the partial and full vaccination. Specifically, the average VPR of the partial vaccination decreased from 57% (SE: 2%) in the pre-Delta period to 40% (SE: 2%) in the Delta-dominated period, suggesting that only the partial shot was insufficient to protect against the Delta variant. Despite the Delta-variant also reduced the average VPR of the full dose from 82% (SE: 4%) in the pre-Delta period to 61% (SE:3%) in the Delta-dominated period, the average VPR of the full vaccination still stayed above the WHO recognized 50% level of vaccine efficacy in most countries except Turkey (Figure 1). The coming of Omicron had reduced the VPRs of the partial and full vaccination in the seven countries to less than 50%. In the Intervening II period, the VPRs were 5.5%-34% (Average: 22.2%, SE: 4.0%) for partial vaccination, and 37%-56% (Average: 49.1%, SE: 2.3%) for full vaccination. When the Omicron variant became prevalent, VPRs were even lower, which were 3.8%-28.5% (Average: 11.5%, SE: 3.3%) and 26%-45% (Average: 38.6%, SE: 2.4%) for the partial and full vaccination, respectively.

**Figure 1:**
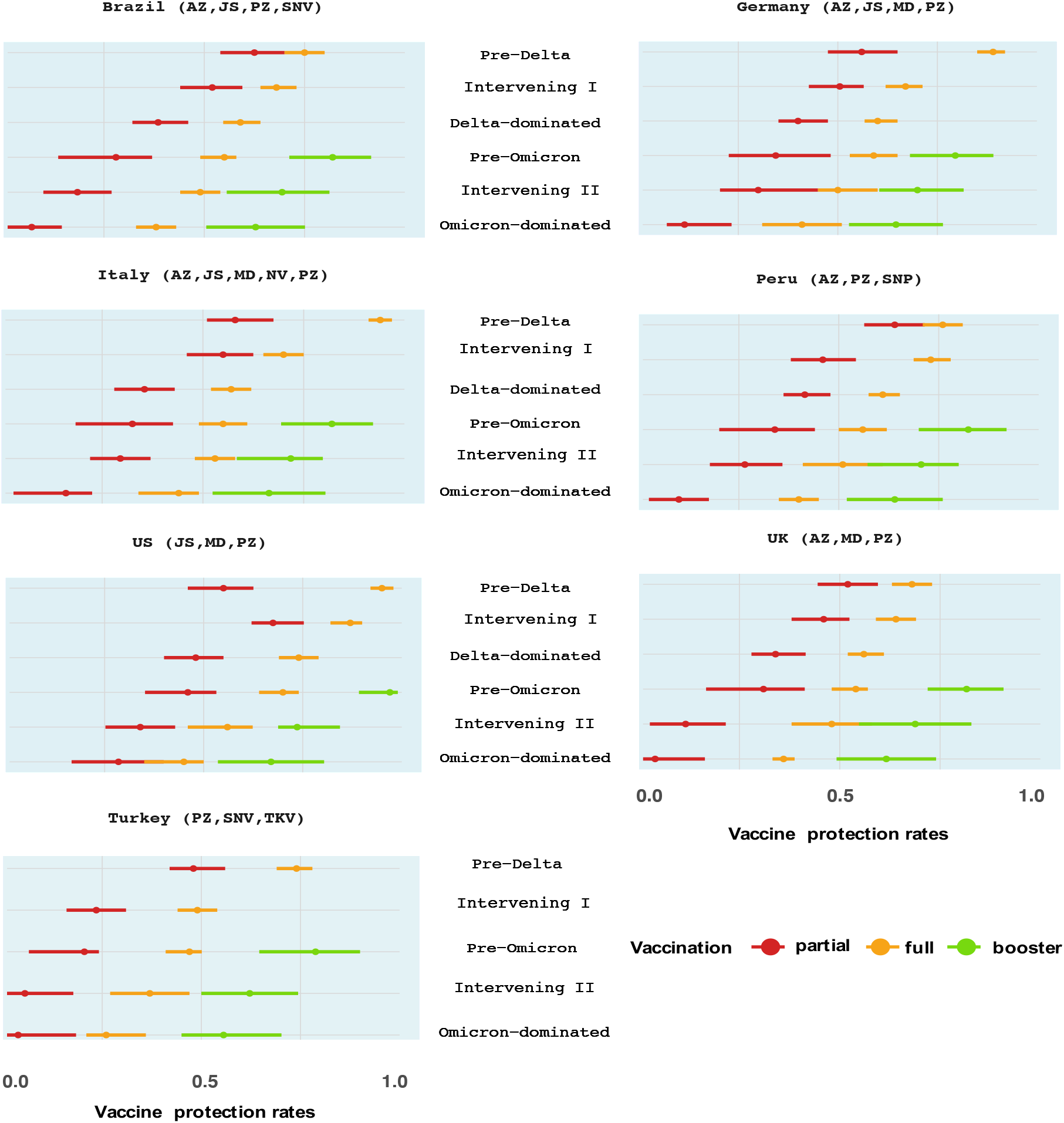
Estimated vaccine protection rates of partial, full and booster vaccination in the seven countries over the six periods with the 95% confidence interval bars. The vaccines used are reported in the parentheses (AZ: AstraZeneca, JS: Janssen, MD: Moderna, NV: Novavax, PZ: Pfizer, SNP: Sinopharm, SNV: Sinovac, TKV: Turkovac).

The booster shot was started in the pre-Omicron period when the Delta was dominant. Our study shows that it readily restored the VPRs to 78.8%-97% (Average: 83.3%, SE: 2.3%), which means that the booster vaccination’s VPRs were 20.5%-31.8% (Average: 26.8%, SE: 1.2%) higher than those of the full vaccination against the Delta variant. In the Omicron-dominated period, the booster shot’s VPRs ranged 55.6%-67.0% (Average: 63%, SE: 1.4%), largely stayed above the 50% threshold. These suggest that the booster shot provided enhanced and effective protection against both the Delta and Omicron variants.

### Impacts of Full and Booster Vaccines

To further evaluate the protection of the COVID-19 vaccination, we investigate the impacts of the full and the booster vaccination on the size of the epidemics and deaths. Five vaccination scenarios were designed: (i) no vaccination at all; (ii) receiving the partial but no full vaccination; (iii) receiving the partial and full vaccination but no booster shots; receiving the booster shots only at the half (iv) and twice (v) of the actual daily booster coverage rates. The impacts of these scenarios were projected using the stochastic epidemic model with the estimated parameters for each country. See the specific designs of the scenario analysis (SA) in Section S5 of the supplementary material (SM).

The projected cumulative confirmed cases and deaths from the start of vaccination to the start of boosters for the seven countries under the no vaccination (i) and only the partial vaccination (ii) scenarios are shown in Figure 2 (a) and (b) with the detailed numerical values listed in Table S4. It shows that no vaccination at all would bring, respectively, 112918 (CI: 96142-129695, Percentage: 242%) and 825 (CI: 630-1020, Percentage: 83%) thousands increase in the cumulative confirmed cases and deaths in the seven countries relative to the observed values under the actual vaccination arrangement. Under the only partial vaccination, the cumulative confirmed cases and deaths would increase by 39697 (CI: 30105-49289, Percentage: 85%) and 218 (CI: 124-312, Percentage: 22%) thousands, respectively, in the seven countries. These two scenario analyses show the significant benefit of the partial and the full vaccination.

**Figure 2:**
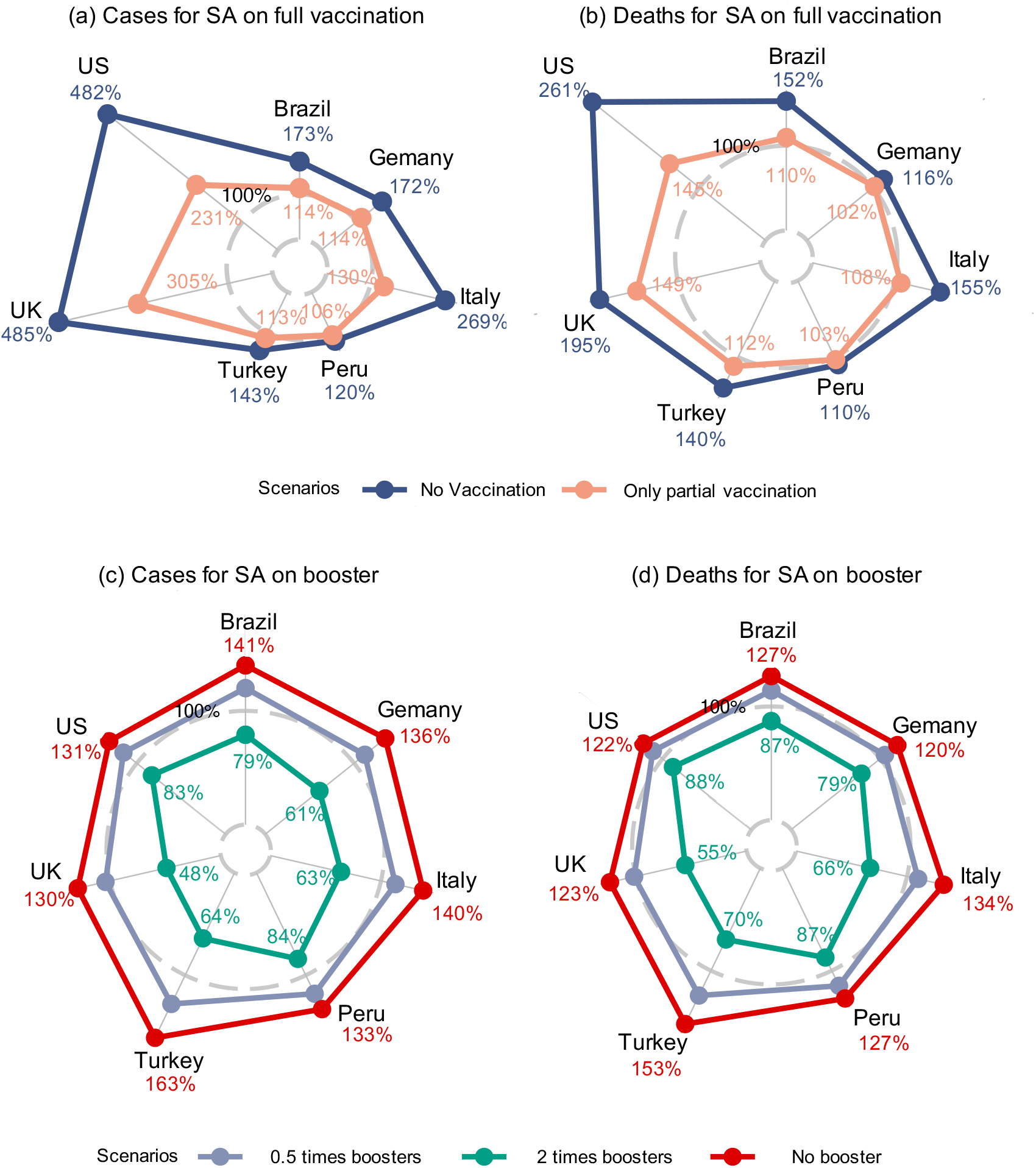
Radar plots on the proportions of the projected cumulative numbers of confirmed cases and deaths under two sets of scenario analyses (SA): the pre-booster vaccine periods under no and partial vaccination (a, b) and the post-booster periods under the no, half and twice booster up-take scenarios (c, d), relative to their respective observed values in the seven countries. The 100% gray dashed circles represent the observed situations.

The less amount of increase in the confirmed cases and deaths under the no and partial vaccination scenarios in Peru was due to its low and slow pace of vaccination, with only 6% and 3% of the population having received the partial and full vaccination within the first 100 days of vaccination. In contrast, 13.4%-18.9% and 5.5%-13.9% of the populations had been partially and fully vaccinated in Germany, Italy, Brazil and Turkey, and the US and UK had the highest vaccination rates of 27.7%-48.1% and 15.5%-15.9% for the partial and full vaccination over the same period (Figure 3 (a)). That US and UK had the highest vaccination rates in the first 100 days led to much higher numbers of cases and deaths under the no and partial vaccination scenarios in Figure 2 (a) and (b), as compared with the other countries.

**Figure 3:**
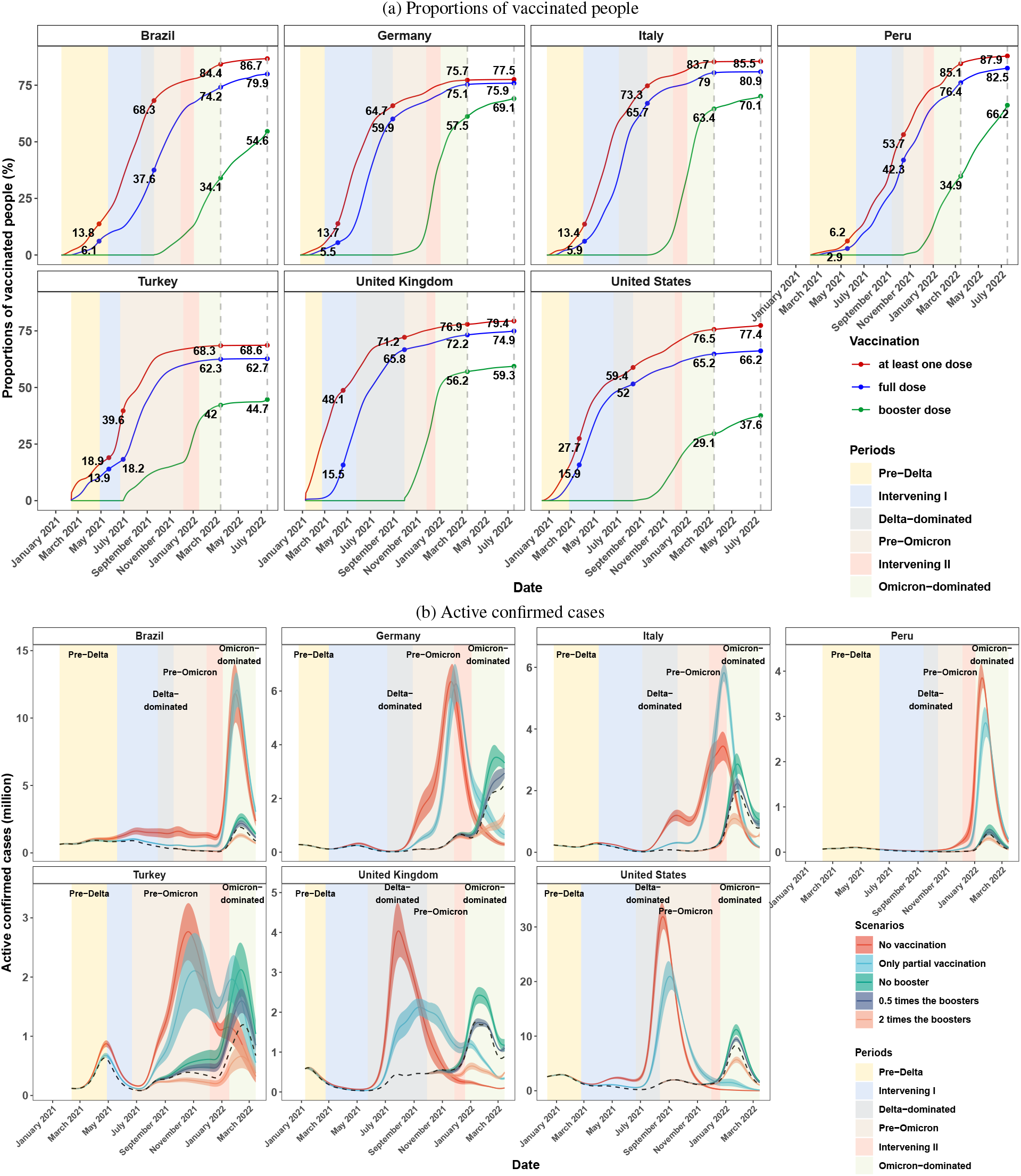
(a) Daily population proportions receiving at least one (red), full (blue) and booster (green) dose of vaccines. The dashed vertical lines mark March 15 and July 17, 2022. (b) The actual (black dashed lines), and the projected daily numbers (in millions) of active confirmed cases (color curves) and their 95% confidence bands (color area) under the five vaccination scenarios.

Figure 2 (c) and (d) display the projected cumulative confirmed cases and deaths from the start of booster shots to March 15, 2022 under the scenarios (iii)-(v) regarding the booster vaccination which kept the observed numbers of the partial and full doses as the baselines; see Table S5 for the detailed numerical values. It is shown that during the post-booster periods, not having the booster shots at all would mean 34860 (CI: 23543-46177) and 143 (CI: 88-198) thousands more confirmed cases and deaths, respectively, in the seven countries, amounting to 36% and 26% increases in the total confirmed cases and deaths, respectively. In the half-booster scenario, the increases in the confirmed cases and deaths would be less than those under the no-booster case, but still translate to 14587 (CI: 8024-21150, Percentage: 15%) and 66 (CI: 29-103, Percentage: 12%) thousands more confirmed cases and death relative to the observed numbers, respectively, for the seven countries in the post-booster period.

If the booster taking rates were doubled, we see decreases of 27679 (CI: 21234-34124, Percentage: 29%) and 94 (CI: 62-126, Percentage: 17%) thousands in total confirmed cases and deaths for the seven countries in the post-booster period. It is noted that the relatively large reductions in the confirmed cases and deaths in Germany, Italy, Turkey and UK under the double booster scenario were due to their actual higher (more than 40%) rates of taking the booster shots by March 15, 2022 (Figure 3 (a)). In contrast, the US, Brazil, Peru had lower booster taking rates, which led to smaller amount of reductions in the confirmed cases and deaths as compared to the other countries.

To further evaluate the dynamics of the COVID-19 epidemics with respect to different vaccination strategies, we report in Figure 3 (b) the observed and the projected daily numbers of active confirmed cases (those confirmed infective people who have not recovered or died), to reflect the potential real-time demand on the hospital system under the five scenarios. It shows that, compared to the observed time series of the active confirmed cases, the peaks of the active confirmed cases would be much elevated and happen much earlier under the no and partial vaccination scenarios. In particular, the numbers of active confirmed cases in Germany, Italy, Turkey, the UK and the US would peak when the Delta was dominant. It also shows that the full and the booster shots significantly delayed the timing and flattened the magnitude of the infection peaks in the seven countries, and in particular, protected the populations in the more lethal pre-Omicron era. As shown in Table S6, the projected numbers of active confirmed cases would exceed the observed peaks for 70-111, 65-156, 23-59 and 0-42 days with the projected peaks being 1.7-9.4 (Mean 4.1, SE 1.1), 1.2-7.0 (Mean 3.5, SE 0.9), 1.2-1.8 (Mean 1.4, SE 0.06) and 1.0-1.3 (Mean 1.1, SE 0.04) times the observed peaks under the no vaccination, partial vaccination, no booster and half booster scenarios, respectively. Those indicate the effect of the vaccination potentially avoided severe runs on the health system of the seven countries.

Comparing the actual observations with the three scenarios regarding the booster shot taking, the peak values of active confirmed people in the seven countries would increase by 23%-78% (Mean: 43%, SE: 6%) under the no booster scenario, and decrease by 26%-63% (Mean: 41%, SE: 5%) relative to the observed peaks under the twice booster scenario, which verifies the booster does can further relieve the pressure on the healthcare system in the Omicron era due to its higher VPRs against the Omicron variants as reported in Figure 1.

It is noted that in Italy the projected peak under the partial vaccination scenario was higher than that under the no vaccination in the Intervening II period. This was due to that a considerable proportion of population would have been infected in the Delta-dominated period under the no vaccine scenario, and Italy had the highest rate of partial vaccination before the Intervening II period among the seven countries (Figure 3 (a)). However, the immunity acquired from the partial vaccination gradually expired without further injected immunity from the full dose. This would lead to a rebound in the numbers of susceptibles, even exceeding those under the no vaccination scenario as shown in Figure S1. This result suggests the importance of acquiring additional immunity through full and booster doses.

## Discussion

This study targets the population protection rates of vaccines. Although the vaccine efficacies are different between different age groups, our results reveal the overall protection rates of a country, which are informative on the total infection size and the demand on the health resources of a country. It is shown that the real-world vaccine protection rates (VPRs) of the partial, full and booster vaccination decreased with time. The full vaccination was effective before Omicron with the VPRs stayed above 50%, which became insufficient when the Omicron was dominant. The booster shot was effective in slowing down the epidemics in both the Delta and Omicron-dominated periods with the average VPRs well above the 50% threshold. Our results on the real-world VPRs were consistent with the vaccine effectiveness in the existing cohort or case-control studies ^9, 12, 16^. The necessity of the full and the booster vaccination are further highlighted by significant reductions in daily numbers of active confirmed cases in the scenario analyses.

Two sets of sensitivity analyses have been conducted to explore the impact of the uncertainty associated with model parameters for the daily asymptomatic rate 1 − *θ*_*t*_ and the average time duration *µ*_*r*_ from recovery to loss of natural immunity (average duration for reinfection) on the estimated VPRs. The sensitivity analyses show that the average absolute differences between the VPRs by using different values of the daily asymptomatic rate and the average duration for reinfection were 1.26% (SE: 0.23%) and 0.94% (SE: 0.17%), respectively, indicating the robustness of the estimated VPRs with respect to the two key parameters.

Despite the effectiveness of the booster shot in the Omicron era, the booster vaccine coverage had a rather slow pace of growth with less than 9% increase from March 15 2022 to July 17 2022 in Italy, Turkey, the UK and the US, as shown in Figure 3 (a). The booster coverage was 37.6% on July 17, 2022 in the US, which was only increased by 8.5% over the four months since March 15 2022, and the UK’s increased only 3.1% over the same period. Thus, there is ample room for vigorous promotion of booster shots in all countries to realize their benefits in reducing both the size of the epidemics and death. The encouraging effects of the booster shots also encourage consideration for another dose after the booster shot to cope with the continuing evolution of the SARS-Cov-2 viruses.

## Methods

The study period in this research is from the start of the vaccine in a country to March 15, 2022, while part of the pre-vaccine period was considered for model parameter estimation. For each country, we divide the post-vaccine era into six consecutive non-overlapping periods: the pre-Delta period from the start of vaccination till the Delta variant was first detected in the country, the following intervening period (Intervening I) until the Delta variant became predominant (more than 50% of the daily detected cases), the Delta-dominated period when the majority of the cases were caused by the Delta variant till the start of booster shots, the pre-Omicron period from the start of booster shots till the Omicron variant was first detected, the intervening period (Intervening II) till Omicron became predominant, the Omicron-dominated period when the majority of the cases were caused by the Omicron variant. It is noted that the dominated variant was still Delta in the pre-Omicron period in the seven countries. Since the start of the booster shot (June 21, 2021) was close to the date (June 29, 2021) when the Delta variant began to dominate in Turkey, we merged its Delta-dominated period and pre-Omicron period, which makes Turkey have only five periods. The key dates of these periods are provided in Table S2 in the SM.

We estimate the real-world VPR by building a stochastic epidemiological model with eleven-compartments to quantify the epidemic process and developing a novel estimation procedure for its parameters.

### Stochastic epidemiological model

The stochastic epidemiological model describes the spread of the SARS-CoV-2 virus and the daily increase of infected cases in a country. The compartments and flows between compartments are shown in Figure 4. The model allows non-permanent vaccine and natural immunity, breakthroughs in vaccinated people, and being asymptomatic and infectious before clinical diagnosis (pre-symptomatic). The susceptible population are divided into five compartments, the ones with no, partial, full and booster vaccine immunity and the ones who have been vaccinated but lost vaccine immunity or have recovered from previous infection but lost natural immunity, in the top row of Figure 4. The currently uninfected compartment without vaccine or natural immunity consists of unvaccinated people, the vaccinated people with expired vaccine immunity and the recovered with expired natural immunity; see the SM for the specific arrangement on the vaccine expiration parameters *µ*_1_, *µ*_2_ and *µ*_3_, and the parameter of losing natural immunity *µ*_*r*_. The latter is responsible for reinfection. The uninfected individuals may catch the virus by contacting with the infected which are divided into three compartments: asymptomatic, pre-symptomatic and diagnosed. Asymptomatic cases represent the ones show no symptoms and do not take a test. The pre-symptomatic period stands for the period after infection but before lab confirmation. The pre-symptomatic cases would be diagnosed at the rate *α* ∈ (0, 1), where 1*/α* represents the average days between being infected and lab diagnosis.

**Figure 4:**
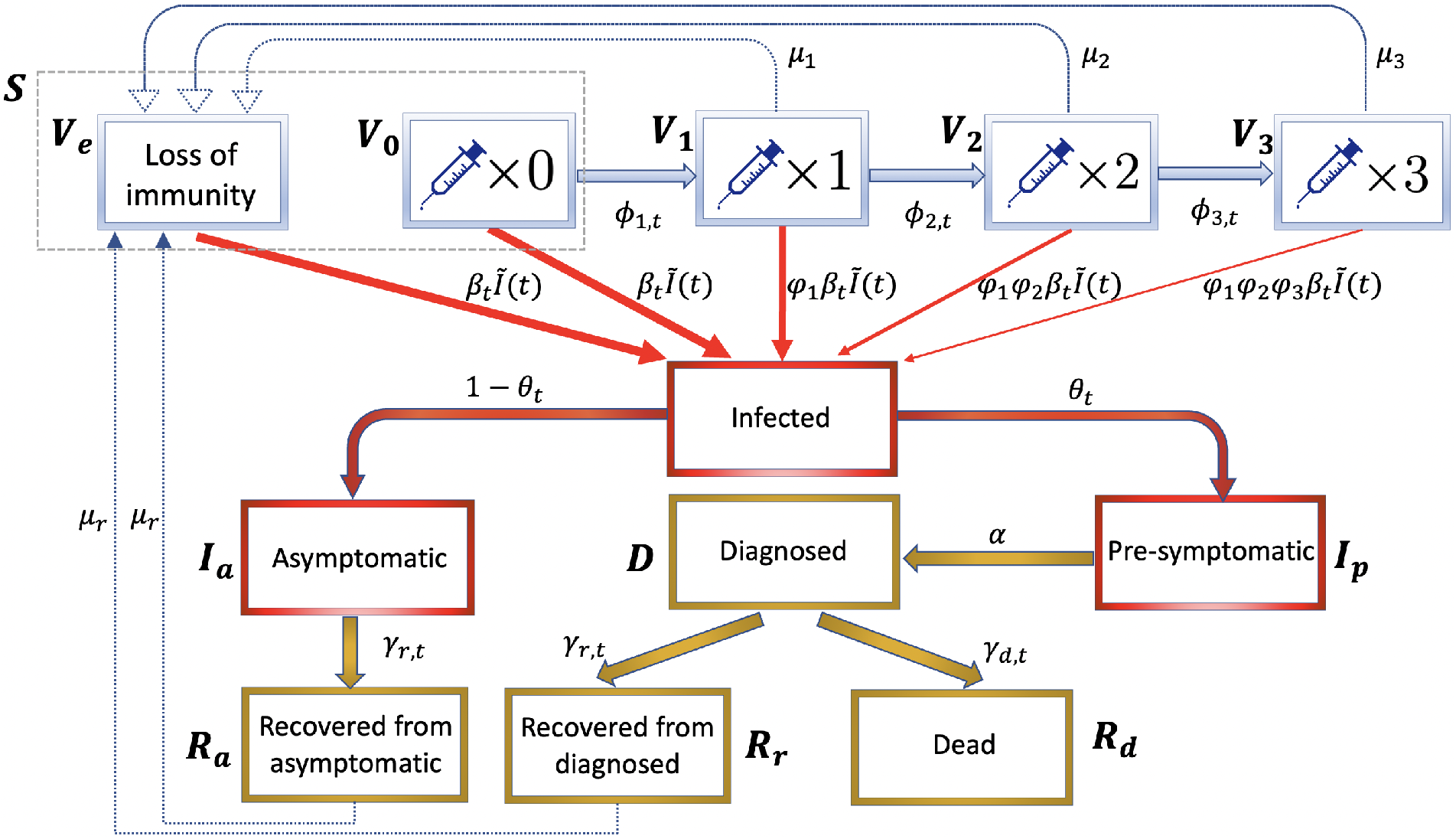
Compartments and their structure flows of the proposed epidemiological model, where the compartments *V*_0_ and *V*_*e*_ constitute state *S* = *V*_0_ + *V*_*e*_ of currently uninfected people without immunity, and *φ*_1_, *φ*_2_ and *φ*_3_ are the parameters representing vaccine protection rates.

Following the set up of the infection rates for different stages of infections^17^, we assume the pre-symptomatic compartment is five times more infectious than the asymptomatic and diagnosed compartments, as diagnosed cases would take precautions and quarantine at home, and asymptomatic cases have no symptoms and should be less infectious. Let *M* be the total population size, *β*_*t*_ denote the time-varying infection rate of the pre-symptomatic cases, and *Ĩ*(*t*) be the standardized total infection load, which is equal to the size of the pre-symptomatic compartment divided by *M* plus those of the asymptomatic and diagnosed compartments divided by 5*M*. Allowing time-varying infection rate (*β*_*t*_) is needed as government and citizens’ responses to COVID-19 change over time and the virus itself keeps mutating. We assume the daily new infections from the susceptible groups without vaccine or natural immunity, and with partial, full and booster vaccine immunity follow conditional Poisson distributions with means equal to *β*_*t*_*Ĩ*(*t*), *φ*_1_*β*_*t*_*Ĩ*(*t*), *φ*_1_*φ*_2_*β*_*t*_*Ĩ*(*t*) and *φ*_1_*φ*_2_*φ*_3_*β*_*t*_*Ĩ*(*t*) multiplying the size of the corresponding group, respectively, which are shown in Figure 4. And a new infection has 1 − *θ*_*t*_ probability being asymptomatic, which is modelled by a binomial distribution. The specification of *θ*_*t*_ is given in the SM, which is based on existing studies^18–20^ on the proportion of asymptomatic cases over different periods of the pandemic. The vaccine effects are reflected by *φ*_1_, *φ*_2_, *φ*_3_ ∈ (0, 1), where the individuals having partial, full and booster vaccine immunity are less likely being infected comparing to the those without vaccine immunity by the factors *φ*_1_, *φ*_1_*φ*_2_ and *φ*_1_*φ*_2_*φ*_3_, respectively. The VPRs for the three vaccine compartments are 1 − *φ*_1_, 1 − *φ*_1_*φ*_2_ and 1 − *φ*_1_*φ*_2_*φ*_3_, respectively.

### Estimation

Given the above model arrangement, we estimate the diagnosis rate *α*, time-varying infection, recovery and death rates, and the VPR parameters *φ*_1_, *φ*_2_ and *φ*_3_ of a country by a multi-step multi-time range procedure via minimizing certain criterion functions using different periods of data for different variants of SARS-CoV-2 virus, and a nonparametric regression method for the time-varying parameters.

The recovery and death rates are estimated by the kernel smoothing regression of daily new recovery and death numbers on the total size of active confirmed infections at time *t*. If the recovery data are not available, we set it as 1*/*14, meaning 14 days as the average recovery time since diagnosis^21^. Let *I*_*p*_(*t*) be the size of the pre-symptomatic compartment and Δ*N* (*t*) be the new confirmed cases at day *t*. Adopting a multi-step estimation procedure, we use the B-splines to fit the infection rate *β*_t_, and minimize the criteria function 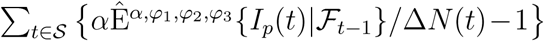^2^ with respect to *α, φ*_1_, *φ*_2_, *φ*_3_ and the coefficients of the B-splines over an estimation period S. Here Δ*N* (*t*)*/α* stands for an imputation for *I*_*p*_(*t*) and 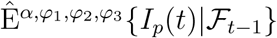 is a simulation-based estimate of the conditional expectation of *I*_*p*_(*t*) given data up to *t* − 1 by averaging the simulated trajectories under the proposed model at each *α, φ*_1_, *φ*_2_, *φ*_3_ and *β*_*t*_. This estimation approach can be viewed as a minimum distance method that minimizes the distance between the trajectories implied by the model and the observed data of daily new confirmed cases. However, due to the unobservable compartments, imputation is needed as well as a decentralized strategy that estimates different parameters over different time periods. The mathematical expressions of the proposed model and the detailed estimation procedure are provided in the SM.

### Sensitivity analysis

The number of reported cases are influenced by the severity of symptoms, public willingness to do testing and the testing capacity. These factors determine the pre-symptomatic proportion *θ*_*t*_ and the diagnosis rate *α* of the pre-symptomatic cases in our model. While *α* is empirically estimated by the proposed method, *θ*_*t*_ is determined from existing studies. Murray et al. (2022)^20^ suggested the asymptomatic cases accounted for 80-90% of COVID-19 infections in the Omicron era. In the main analysis, the probability for new infections being asymptomatic since the detection of Omicron variant was assumed to increase linearly from 40% to 90%. To explore the sensitivity of this specification, we conducted a sensitivity analysis which assumed the probability of being asymptomatic for new infections after the Omicron increased linearly from 40% to 80%. The estimated VPRs in the intervening II and the Omicron-dominated periods for the seven countries are reported in Table S7, which shows that the magnitude of the differences between VPRs by altering the daily asymptomatic rates 1 − *θ*_*t*_ was no more than 8.6% and the average of the absolute differences was 1.26% (SE: 0.23%). We have also conducted a sensitivity analysis on the estimated VPRs with respect to *µ*_*r*_, the time duration from recovery to loss of natural immunity. It shows that the differences between the estimated VPRs in the seven countries with *µ*_*r*_ being 16 months as in the main analysis and those with *µ*_*r*_ being 6 months were at most 5% apart with the average absolute differences being 0.94% (SE: 0.17%). Table S7 contains the details for the two sets of sensitivity analyses.

### Reporting summary

Further information on research design is available in the Nature Research Reporting summary linked to this article.

## Supporting information

This supplementary material provides the additional details, tables and figures to the main paper.

## Data Availability

All data produced are available online at "https://covid.ourworldindata.org/".

## Data availability

The daily epidemiological and vaccination data that support the findings of this study are available in a public repository “Our World in Data”, [“https://covid.ourworldindata.org/”].

## Code availability

The codes used in this study are available on GitHub at https://github.com/zyrstat/Estimating-VPRs.

## Author contributions

Y.Z., Y.Q, and S.X.C designed the study and drafted the paper. Y.Z. processed the data and performed the statistical analysis. Y.Z., Y.Q, and S.X.C interpreted the results of the analysis and reviewed the paper. All authors revised the paper and gave final approval for the version to be submitted.

