## Supplementary material for "Real-World COVID-19 Vaccine Protection Rates against Infection in the Delta and Omicron Eras": This supplementary material provides the additional details, tables and figures to the main paper.

**Corresponding authors.*

This supplementary material provides the additional details, tables and figures to the main paper.

### S1 Data

We obtained the publicly available nation-wide epidemiological and vaccination data from February 23, 2020 to March 15, 2022, for the 7 countries considered in this study. The daily cumulative numbers of the confirmed cases and deaths were obtained from “the 2019 Novel Coronavirus Visual Dashboard” at Johns Hopkins University. The daily recovered cases were imputed using 14 days as the average time of recovery from diagnosis by Equation (S.3), since recoveries have not been reported since August for those countries. The information on the vaccine types and the cumulative numbers of people having received the partial, full and booster vaccination were obtained from the official reports of the countries[1]. To reduce measurement errors, we used the kernel smoothing approach[2] to smooth the daily observed data before analysis.

### S2 Epidemiological model

In this section, we provide detailed explanations for the proposed epidemic model described in Figure 4. Let $V_0(t)$, $V_1(t)$, $V_2(t)$, $V_3(t)$ and $V_e(t)$ be the counts of five uninfected sub-populations having received no vaccine, with partial, full, booster vaccine immunity, and with expired vaccine or natural immunity at date $t$, respectively. We combine $V_0$ and $V_e$ into one state $S$, and let $S(t) = V_0(t) + V_e(t)$ be the counts of currently uninfected people without vaccine or natural immunity whether due to receiving no vaccine or losing immunity from vaccines or previous infections at day $t$. Let $\phi_{1,t}$, $\phi_{2,t}$ and $\phi_{3,t}$ be the time-varying vaccination rates from $V_0(t)$ to $V_1(t)$, $V_1(t)$ to $V_2(t)$, and $V_2(t)$ to $V_3(t)$, respectively. We consider the temporary vaccine-induced immunity and natural immunity with the average lengths of immunity after the partial, full and booster doses of vaccines and the recovery from previous infection being $1/\mu_1$, $1/\mu_2$, $1/\mu_3$ and $1/\mu_r$ days, respectively. We set $\mu_1 = 1/60$, $\mu_2 = 1/240$, $\mu_3 = 1/300$ and $\mu_1 = 1/56$, $\mu_2 = 1/90$, $\mu_3 = 1/140$ before and after the emergence of the Omicron variant according to the existing studies[3–8]. Motivated by the study which shows that SARS-CoV-2 reinfections were uncommon (less than 1% of the total confirmed infections) until the end of 2021[9], we set $\mu_r = 0$ before the emergence of the Omicron variant, and set $\mu_r = 1/480$ after the emergence of the Omicron variant in the main analysis and $\mu_r = 1/180$ in the sensitivity analysis based on studies[10–12] on the duration of immune protection from infection.

Let $I_a(t)$, $I_p(t)$ and $D(t)$ be the counts for the asymptomatic, pre-symptomatic and diagnosed compartments with the time-varying infection rates $\beta_t^{\mathrm{I}_a}$, $\beta_t^{\mathrm{I}_p}$ and $\beta_t^{\mathrm{D}}$, respectively. Those are the three infectious states in the proposed model. Following the specification[13], we set the infection rates of the asymptomatic and diagnosed individuals to be 20% of that of symptomatic ones, namely $\beta_t^{\mathrm{I}_a} = \beta_t^{\mathrm{D}} = \beta_t^{\mathrm{I}_p}/\zeta$ with $\zeta = 5$. The likelihoods of being infected in the states $V_1$, $V_2$ and $V_3$ with vaccine immunity are assumed to be $\varphi_1$, $\varphi_1\varphi_2$ and $\varphi_1\varphi_2\varphi_3$ as that in the susceptible state without vaccine or natural immunity $S$, respectively, where $\varphi_1$, $\varphi_2$, and $\varphi_3$ are three constants between 0 and 1. Let $H(t, \boldsymbol{\beta}_t) = \{\beta_t^{\mathrm{I}_a} I_a(t) + \beta_t^{\mathrm{I}_p} I_p(t) + \beta_t^{\mathrm{D}} D(t)\}/M$ be the total infection loading at $t$, where $\boldsymbol{\beta}_t = (\beta_t^{\mathrm{I}_a}, \beta_t^{\mathrm{I}_p}, \beta_t^{\mathrm{D}})^T$. Via contacts with the infectious states, the currently uninfected people are infected with the intensities $H(t, \boldsymbol{\beta}_t)S(t)$, $\varphi_1 H(t, \boldsymbol{\beta}_t)V_1(t)$, $\varphi_1\varphi_2 H(t, \boldsymbol{\beta}_t)V_2(t)$ and $\varphi_1\varphi_2\varphi_3 H(t, \boldsymbol{\beta}_t)V_3(t)$ for the states $S$, $V_1$, $V_2$ and $V_3$, respectively.

All infected people will develop either asymptomatic $I_a$ or symptomatic $I_p$. Recall that $\theta_t$ denotes the daily proportion of being pre-symptomatic. As existing studies found 20% of COVID-19 infections were asymptomatic at the early stage of the COVID-19 pandemic[14], 40% before the detection of Omicron[15], and 80–90% for Omicron[16], we assume $\theta_t$ to be a piecewise linear function, which is 0.8 till the first detection of the Delta variant, then linearly decreases to 0.6 till the first detection of the Omicron variant, and then till March 15, 2022, linearly decreases to 0.1 in the main analysis and to 0.2 in the sensitivity analysis. All asymptomatic infections are never diagnosed and will recover naturally $R_a$ with the recovery rate $\gamma_{r,t}$. The pre-symptomatic cases will be confirmed and moved to the diagnosed state $D$ with the diagnosis rate $\alpha$ in a future date, then to the recovered $R_r$ with the recovery rate $\gamma_{r,t}$ and the dead $R_d$ with the death rate $\gamma_{d,t}$. Let

$\Delta$ denote the daily change of a compartment. Recall that $S(t) = V_0(t) + V_e(t)$. The following equations present the relationship of the conditional means of the aforementioned compartments at time $t$:

$$
\begin{aligned}
\mathrm{E}\{\Delta V_0(t)|\mathcal{F}_t\} &= -H(t,\boldsymbol{\beta}_t)V_0(t) - \phi_{1,t}V_0(t) \\
\mathrm{E}\{\Delta V_e(t)|\mathcal{F}_t\} &= -H(t,\boldsymbol{\beta}_t)V_e(t) + \mu_1 V_1(t) + \mu_2 V_2(t) + \mu_3 V_3(t) + \mu_r R_r(t) + \mu_r R_a(t), \\
\mathrm{E}\{\Delta V_1(t)|\mathcal{F}_t\} &= \phi_{1,t}V_0(t) - \mu_1 V_1(t) - \varphi_1 H(t,\boldsymbol{\beta}_t)V_1(t) - \phi_{2,t}V_1(t), \\
\mathrm{E}\{\Delta V_2(t)|\mathcal{F}_t\} &= \phi_{2,t}V_1(t) - \mu_2 V_2(t) - \varphi_1\varphi_2 H(t,\boldsymbol{\beta}_t)V_2(t) - \phi_{3,t}V_2(t), \\
\mathrm{E}\{\Delta V_3(t)|\mathcal{F}_t\} &= \phi_{3,t}V_2(t) - \mu_3 V_3(t) - \varphi_1\varphi_2\varphi_3 H(t,\boldsymbol{\beta}_t)V_3(t), \qquad \text{(S.1)} \\
\mathrm{E}\{\Delta I_a(t)|\mathcal{F}_t\} &= (1-\theta_t)H(t,\boldsymbol{\beta}_t)\{S(t) + \varphi_1 V_1(t) + \varphi_1\varphi_2 V_2(t) + \varphi_1\varphi_2\varphi_3 V_3(t)\} - \gamma_{r,t}I_a(t), \\
\mathrm{E}\{\Delta I_p(t)|\mathcal{F}_t\} &= \theta_t H(t,\boldsymbol{\beta}_t)\{S(t) + \varphi_1 V_1(t) + \varphi_1\varphi_2 V_2(t) + \varphi_1\varphi_2\varphi_3 V_3(t)\} - \alpha I_p(t), \\
\mathrm{E}\{\Delta D(t)|\mathcal{F}_t\} &= \alpha I_p(t) - (\gamma_{r,t} + \gamma_{d,t})D(t),\ \mathrm{E}\{\Delta R_a(t)|\mathcal{F}_t\} = \gamma_{r,t}I_a(t) - \mu_r R_a(t), \\
\mathrm{E}\{\Delta R_r(t)|\mathcal{F}_t\} &= \gamma_{r,t}D(t) - \mu_r R_r(t) \text{ and } \mathrm{E}\{\Delta R_d(t)|\mathcal{F}_t\} = \gamma_{d,t}D(t).
\end{aligned}
$$

The proposed stochastic epidemic model assumes the daily increments of those compartments follow Poisson distributions with the conditional means specified by the equations in (S.1). It can be shown that the effective reproduction number $R_t$ is

$$R_t = \left\{(1-\theta_t)\frac{\beta_t^{\mathrm{I}_a}}{\gamma_{r,t}} + \theta_t\left(\frac{\beta_t^{\mathrm{I}_p}}{\alpha} + \frac{\beta_t^{\mathrm{D}}}{\gamma_t}\right)\right\}\frac{S(t) + \varphi_1 V_1(t) + \varphi_1\varphi_2 V_2(t) + \varphi_1\varphi_2\varphi_3 V_3(t)}{M} \qquad \text{(S.2)}$$

under the proposed model, where $\gamma_t = \gamma_{r,t} + \gamma_{d,t}$. The effective reproduction number is a key epidemiological parameter. When $R_t > 1 (< 1)$, the epidemic is increasing (decreasing).

### S3 Estimation

Note that the observed data of a country are $\{N(t), R_d(t), G_1(t), G_2(t), G_3(t)\}_{t=1}^{T}$, where $N(t)$, $R_d(t)$, $G_1(t)$, $G_2(t)$ and $G_3(t)$ are the daily cumulative numbers of the confirmed cases, deaths, the partial, full and booster vaccinated people. Since the recovery data were only reported at the beginning of the pandemic, with the recovery rate $\gamma_{r,t}$ set as $1/14$, we impute the numbers of recovery (when it was not available) and active confirmed infections by

$$\begin{aligned} \hat{R}_r(t) &= \hat{R}_r(t-1) + \hat{D}(t-1)/14 - \mu_r \hat{R}_r(t-1), \\ \hat{D}(t) &= \hat{D}(t-1) + \Delta N(t-1) - \hat{D}(t-1)/14 - \Delta R_d(t-1). \end{aligned} \tag{S.3}$$

Following the multi-step decentralized estimation procedure developed in Zhu et al. (2022)[17], the steps below summarize the main steps for estimating the parameters $\alpha$, $\beta_t^{\mathrm{I}_p}$, $\gamma_{d,t}$, $\gamma_{r,t}$, $\varphi_1$, $\varphi_2$ and $\varphi_3$ in the proposed model, which lead to the estimation of the effective reproduction number $R_t$ via the Equation (S.2) and the vaccine protection rates $1-\varphi_1$, $1-\varphi_1\varphi_2$, $1-\varphi_1\varphi_2\varphi_3$ for the partial, full and booster shots of a country.

1. *Estimation of removal rates $\gamma_{d,t}$ and $\gamma_{r,t}$.* From the last two equations in (S.1), we estimate $\gamma_{d,t}$ and $\gamma_{r,t}$ by local linear regression of the daily new deaths $\Delta R_d(t)$ and daily new recoveries $\Delta R_r(t)$ on $D(t)$ via

$$\begin{aligned} \hat{\gamma}_{d,t} &= \frac{\sum_{i=1}^{T-1} D(i)\Delta R_d(i) B((t-i)/h_d)}{\sum_{i=1}^{T-1} D(i)^2 B((t-i)/h_d)} \quad \text{and} \\ \hat{\gamma}_{r,t} &= \frac{\sum_{i=1}^{T-1} D(i)\Delta R_r(i) B((t-i)/h_r)}{\sum_{i=1}^{T-1} D(i)^2 B((t-i)/h_r)}, \end{aligned} \tag{S.4}$$

where $B(\cdot)$ is a kernel function, and $h_d$ and $h_r$ are the temporal smoothing bandwidths[18].

2. *Estimation of diagnosis rate* $\alpha$. We use the time period $\mathcal{S}_1$ of one month right before the start of public vaccination to estimate the diagnosis rate $\alpha$. In this period, there were no vaccine effects, where $\varphi_1 = \varphi_2 = \varphi_3 = 1$ and $G_1(t) = G_2(t) = G_3(t) = 0$. Given $\alpha$ and $\beta_t^{\mathrm{I}_p}$, we consider a contrast measure between the estimated $I_p(t)$ based on the observed data and that simulated from the proposed model, in the form of

$$f_1(\alpha, \beta_t^{\mathrm{I}_p}) = \frac{1}{|\mathcal{S}_1|} \sum_{t \in \mathcal{S}_1} \left\{ \hat{\mathrm{E}}^{\alpha}\{I_p(t)|\mathcal{F}_{t-1}\}/\hat{I}_p^{\alpha}(t) - 1 \right\}^2 \text{ for } \hat{I}_p^{\alpha}(t) = \Delta N(t)/\alpha, \tag{S.5}$$

where $\hat{\mathrm{E}}^{\alpha}\{I_p(t)|\mathcal{F}_{t-1}\}$ is the estimated mean of $I_p(t)$ given all the information at $t-1$ by the simulations from the proposed model using the given $\alpha$ and $\beta_t^{\mathrm{I}_p}$. The nonlinear infection rate $\beta_t^{\mathrm{I}_p}$ is approximated by B-spline functions. We minimize this contrast measure $f_1(\alpha, \beta_t^{\mathrm{I}_p})$ with respect to $\alpha$ and the coefficients of B-spline basis functions of $\beta_t^{\mathrm{I}_p}$ by the grid search algorithm.

3. *Estimation of vaccine effects* $\varphi_1$*,* $\varphi_2$ *and* $\varphi_3$. For each of the six periods with different COVID-19 variants (listed in Table S3), we estimate the VPR parameters $\varphi_1$, $\varphi_2$ and $\varphi_3$ by a similar method as the estimation of $\alpha$. Note that we set $\varphi_3 = 1$ for the periods before the start of booster shot vaccination. Similar to the objective function $f_1(\alpha, \beta_t^{\mathrm{I}_p})$ in (S.5), we consider to minimize

$$f_2(\varphi_1, \varphi_2, \varphi_3, \beta_t^{\mathrm{I}_p}) = \frac{1}{|\mathcal{S}_2|} \sum_{t \in \mathcal{S}_2} \left\{ \hat{\mathrm{E}}^{\hat{\alpha},\varphi_1,\varphi_2,\varphi_3}\{I_p(t)|\mathcal{F}_{t-1}\}/\hat{I}_p^{\hat{\alpha}}(t) - 1 \right\}^2, \tag{S.6}$$

over a time range $\mathcal{S}_2$, where $\hat{I}_p^{\hat{\alpha}}(t) = \Delta N(t)/\hat{\alpha}$ is the imputed value of $I_p(t)$ by the estimated diagnosis rate $\hat{\alpha}$ obtained in the previous step, and $\hat{\mathrm{E}}^{\hat{\alpha},\varphi_1,\varphi_2,\varphi_3}\{I_p(t)|\mathcal{F}_{t-1}\}$ is the estimated expectation of $I_p(t)$ by the simulations from the proposed model using the given parameters. The estimates of $\varphi_1$, $\varphi_2$ and $\varphi_3$ are obtained by minimizing $f_2(\varphi_1, \varphi_2, \varphi_3, \beta_t^{\mathrm{I}_p})$ via the grid search algorithm and B-spline approximation of $\beta_t^{\mathrm{I}_p}$.

4. *Estimation of infection rate $\beta_t^{I_p}$.* As the B-spline estimate of $\beta_t^{\mathrm{I}_p}$ via optimizing the objective functions may not be continuous between the six study periods, we use kernel smoothing method for estimating $\beta_t^{\mathrm{I}_p}$ after obtaining the estimates of $\alpha$, $\varphi_1$, $\varphi_2$ and $\varphi_3$, which is in a similar manner as the smoothing estimates $\hat{\gamma}_{d,t}$ and $\hat{\gamma}_{r,t}$ in (S.4).

Note that $\hat{I}_p^{\hat{\alpha}}(t) = \Delta N(t)/\hat{\alpha}$ is the imputed value of $I_p(t)$. We can impute $I_a(t)$ and $R_a(t)$ as

$$\begin{aligned}\hat{I}_a^{\alpha}(t) &= \{\Delta\hat{I}_p^{\alpha}(t-1) + \alpha\hat{I}_p^{\alpha}(t-1)\}(1-\theta_{t-1})/\theta_{t-1} + (1-\hat{\gamma}_{r,t-1})\hat{I}_a^{\alpha}(t-1) \text{ and} \\ \hat{R}_a^{\alpha}(t) &= \hat{R}_a^{\alpha}(t-1) + \hat{\gamma}_{r,t-1}\hat{I}_a^{\alpha}(t-1) - \mu_r\hat{R}_a^{\hat{\alpha}}(t-1),\end{aligned}$$

respectively. As $\{\Delta I_p(t)+\alpha I_p(t)\}/[\theta_t\{S(t)+\varphi_1 V_1(t)+\varphi_1\varphi_2 V_2(t)+\varphi_1\varphi_2\varphi_3\kappa V_3(t)\}]$ serves as a substitution for the total infection loading $H(t, \boldsymbol{\beta}_t)$, we can impute $V_1(t)$, $V_2(t)$, $V_3(t)$ and $S(t)$ by

$$\begin{aligned}\hat{V}_1^{\hat{\alpha}}(t) =&(1-\mu_1)\hat{V}_1^{\hat{\alpha}}(t-1) + \Delta G_1(t-1) - \Delta G_2(t-1) - \hat{r}(t-1)\hat{\varphi}_1\hat{V}_1^{\hat{\alpha}}(t-1), \\ \hat{V}_2^{\hat{\alpha}}(t) =&(1-\mu_2)\hat{V}_2^{\hat{\alpha}}(t-1) + \Delta G_2(t-1) - \Delta G_3(t-1) - \hat{r}(t-1)\hat{\varphi}_1\hat{\varphi}_2\hat{V}_2^{\hat{\alpha}}(t-1), \\ \hat{V}_3^{\hat{\alpha}}(t) =&(1-\mu_3)\hat{V}_3^{\hat{\alpha}}(t-1) + \Delta G_3(t-1) - \hat{r}(t-1)\hat{\varphi}_1\hat{\varphi}_2\hat{\varphi}_3\hat{V}_3^{\hat{\alpha}}(t-1), \\ \hat{S}^{\hat{\alpha}}(t) =&\hat{S}^{\hat{\alpha}}(t-1) - \Delta G_1(t-1) - \hat{r}(t-1)\hat{S}^{\hat{\alpha}}(t-1) + \\ &\qquad \mu_1\hat{V}_1^{\hat{\alpha}}(t-1) + \mu_2\hat{V}_2^{\hat{\alpha}}(t-1) + \mu_3\hat{V}_3^{\hat{\alpha}}(t-1) + \mu_r\hat{R}_r(t-1) + \mu_r\hat{R}_a^{\hat{\alpha}}(t-1).\end{aligned}$$

with the initial values of $V_1$, $V_2$ and $V_3$ being zero at the start of the vaccination, where

$$\hat{r}(t) = \frac{\hat{I}_p^{\hat{\alpha}}(t+1) - (1-\hat{\alpha})\hat{I}_p^{\hat{\alpha}}(t)}{\theta_t(\hat{S}^{\hat{\alpha}}(t) + \hat{\varphi}_1\hat{V}_1^{\hat{\alpha}}(t) + \hat{\varphi}_1\hat{\varphi}_2\hat{V}_2^{\hat{\alpha}}(t) + \hat{\varphi}_1\hat{\varphi}_2\hat{\varphi}_3\hat{V}_3^{\hat{\alpha}}(t))}.$$

The infection rate $\beta_t^{\mathrm{I}_p}$ can be estimated by nonparametric regression of $Y(t) = \{\hat{I}_p^{\hat{\alpha}}(t+1) + (\hat{\alpha}-1)\hat{I}_p^{\hat{\alpha}}(t)\}/\theta_t$ on $X(t) = \big[\hat{I}_p^{\hat{\alpha}}(t) + \{D(t) + \hat{I}_a^{\hat{\alpha}}(t)\}/\zeta\big]\big\{\hat{S}^{\hat{\alpha}}(t) + \hat{\varphi}_1\hat{V}_1^{\hat{\alpha}}(t) + \hat{\varphi}_1\hat{\varphi}_2\hat{V}_2^{\hat{\alpha}}(t) + \hat{\varphi}_1\hat{\varphi}_2\hat{\varphi}_3\hat{V}_3^{\hat{\alpha}}(t)\big\}/M$ as

$$\hat{\beta}_t^{\mathrm{I}_p} = \frac{\sum_{i=1}^{T-2} X(i)Y(i)B\{(t-i)/h\}}{\sum_{i=1}^{T-2} X(i)^2 B\{(t-i)/h\}}. \tag{S.7}$$

For pre-vaccine eras, $\beta_t^{\mathrm{I}_p}$ is the same estimated as (S.7) with $\hat{V}_1^{\hat{\alpha}}(t)$, $\hat{V}_2^{\hat{\alpha}}(t)$ and $\hat{V}_3^{\hat{\alpha}}(t)$ setting as zero.

5. *Parametric bootstrap inference.* Bootstrap procedure is used to obtain confidence intervals for the estimated parameters in steps 1–4. Given the estimates $\hat{\varphi}_1$, $\hat{\varphi}_2$ and $\hat{\varphi}_3$ from each of the six post-vaccine periods, and $\hat{\alpha}$, $\hat{\beta}_t^{\mathrm{I}_p}$, $\hat{\gamma}_{r,t}$, $\hat{\gamma}_{d,t}$, we generate bootstrap resampled trajectories based on proposed stochastic epidemic model. All the parameters were re-estimated based on the bootstrap resampled observations. The resampling was replicated for a large number ($B$) of times to obtain $B$ independent bootstrap estimates for the parameter. The sample standard deviation and the 2.5% and 97.5% percentiles of the bootstrap estimates can be used to estimate the standard error and to construct confidence intervals for the estimates obtained in steps 1–4.

### S4 Fitting performance

We use the relative errors between the mean projected numbers of cumulative confirmed cases by 1000 simulations with the estimated parameters and the observed numbers of cumulative confirmed cases to evaluate the fitting performance of our proposed model. The relative errors in the main analysis of the seven countries shown in Figure S3 were not higher than 25%, which reflects our model performs well.

### S5 Scenario analysis

Scenario analysis (SA) is conducted to evaluate the impacts of different vaccination strategies on the confirmed cases and death of the pandemic. Five vaccination scenarios were designed: (i) no vaccination at all; (ii) receiving the partial but no full vaccination; (iii) receiving the partial and full vaccination but no booster shots; receiving the booster shots only at the half (iv) and twice (v) of the actual daily booster vaccination rate.

Specifically, in the no and partial vaccination scenarios (i)-(ii) designed for the effects of the full vaccination, simulations were performed from the start of vaccination to March 15, 2022. During this period, the sub-population having received the full and booster doses of vaccines were set to zero while maintaining the actual numbers for having received partial vaccination in the partial vaccination scenario, and the sub-population receiving at least one dose was set to zero in the no vaccination scenario. Similarly, for the last three scenarios (iii)-(v) designed for evaluating the effects of the booster shot, simulations were performed from the start of the booster vaccination with the daily observed numbers of people having received the partial and full vaccination, while the numbers of people having received the booster doses were set to be zero, half and twice the observed vaccine up-takes, respectively. In the twice booster up-take scenario, the would-be numbers of people having received booster doses were truncated to the numbers of full vaccination at the time if the doubling up exceeds the latter number of the country. Under each of the scenarios, the daily adjusted numbers of vaccinated people and the empirically estimated parameters are plugged into the proposed epidemic model (S.1) to project the would-be dynamics of the pandemic.

### S6 Tables

Table S1: The vaccine efficacy (for clinical trials) or vaccine effectiveness (for observational studies) and corresponding 95% confidence interval (in parentheses) obtained in recent studies. Observational studies were more common in the Omicron era. A "–" indicates the primary vaccination of Janssen is one dose.

(a) Vaccine efficacy or effectiveness of one or two doses.

| Vaccine name (type) | Original strain | | Delta variant | | Omicron variant | |
|---|---|---|---|---|---|---|
| | one dose | two doses | one dose | two doses | one dose | two doses |
| Pfizer (mRNA) | 52 (29.5 - 68.4)[19] | 95 (90.3 - 97.6)[19] | 45.2 (43.3 - 47.1)[20] | 90.9 (89.6 - 92.0)[20] | 42.8 (40.3 - 45.1)[20] | 65.5 (63.9 - 67.0)[20] |
| Moderna (mRNA) | 95.2 (91.2 - 97.4)[21] | 94.1 (89.3 - 96.8)[21] | 60.1 (51.8 - 66.9)[20] | 94.5 (90.5 - 96.9)[20] | 47.9 (43.1 - 52.3)[20] | 75.1 (70.8 - 78.7)[20] |
| Janssen (viral vector) | 66.9 (59.0 - 73.4)[22] | – | 60[23] | – | 24 (18 - 29)[24] | – |
| AstraZeneca (viral vector) | 64.1 ( 50.5 - 73.9)[25] | 62.1 (41.0 - 75.7)[25] | 42.9 (39.8 - 45.9)[20] | 82.8 (74.5 - 88.4)[20] | 17.7 (14.3-21.0)[20] | 48.9 (39.2 - 57.1)[20] |
| | 76.0 (59.3 - 85.9)[26] | 66.7 (57.4 - 74.0)[26] | 30.0 (24.3 - 35.3)[27] | 67.0 (61.3 - 71.8)[27] | | |
| Sinopharm (inactivated virus) | 65.5 (52.0 - 75.1)[28] | 78.1 (64.8 - 86.3)[28] | 13.8 (-60.2 - 54.8)[29] | 59.0 (16.0 - 81.6)[29] | | |
| Sinovac (inactivated virus) | 57.9 (46.4 - 66.9) [30] | 50.7 (36.0 - 62.0) [30] | 13.8 (-60.2 - 54.8)[29] | 59.0 (16.0 - 81.6)[29] | 32.7 (14.4 – 47.6)[31] | 25.1 (14.7–34.3)[31] |

(b) Vaccine effectiveness of booster doses.

| Primary | Booster | Delta variant | Omicron variant |
|---|---|---|---|
| AstraZeneca | Pfizer | 95.4 (95.1 - 95.6)[20] | 62.4 (61.8 - 63.0)[20] |
| AstraZeneca | Moderna | 97.0 (96.7 - 97.3)[20] | 70.1 (69.5 - 70.7)[20] |
| AstraZeneca | AstraZeneca | 82.3 (71.3 - 89.0)[20] | 55.6 (44.4 - 64.6)[20] |
| Pfizer | Pfizer | 95.1 (94.8 - 95.4)[20] | 67.2 (66.5 - 67.8)[20] |
| Pfizer | Moderna | 96.6 (96.0 - 97.1)[20] | 73.9 (73.1 - 74.6)[20] |
| Moderna | Pfizer | 94.7 (89.3 - 97.3)[20] | 64.9 (62.3 - 67.3)[20] |
| Moderna | Moderna | 96.4 (91.4 - 98.5)[20] | 66.3 (63.7 - 68.8)[20] |
| Sinovac | Sinovac | | 51.0 (39.6–60.4)[31] |
| Janssen | Janssen | | 54 (43–63)[24] |
| Sinovac | Pfizer | | 63.6 (62.8 - 64.3)[32] |

Table S2: Start dates of vaccination and boosters, first detection of the Delta and Omicron variants, and dates when Delta and Omicron began to dominate in the seven countries. The Delta and Omicron dominant dates in the 4th and 7th columns were reported as the first dates when the proportions of Delta and Omicron exceeded 50% in all SARS-CoV-2 viruses by genome sequencing.

| Country | Start of vaccination | Start of Delta | Date Delta dominates | Start of booster | Start of Omicron | Date Omicron dominates |
|---|---|---|---|---|---|---|
| Brazil | 2021-01-16 | 2021-05-20 | 2021-08-16 | 2021-09-19 | 2021-11-29 | 2022-01-03 |
| Germany | 2020-12-26 | 2021-03-01 | 2021-07-05 | 2021-08-29 | 2021-11-26 | 2022-01-03 |
| Italy | 2020-12-26 | 2021-04-02 | 2021-07-05 | 2021-09-19 | 2021-11-26 | 2022-01-03 |
| Peru | 2021-02-07 | 2021-06-10 | 2021-09-13 | 2021-10-14 | 2021-12-06 | 2022-01-03 |
| Turkey | 2021-02-11 | 2021-04-28 | 2021-06-21 | 2021-06-29 | 2021-12-06 | 2022-01-17 |
| UK | 2021-01-09 | 2021-02-22 | 2021-05-24 | 2021-09-29 | 2021-11-27 | 2021-12-20 |
| US | 2020-12-12 | 2021-02-23 | 2021-06-21 | 2021-08-12 | 2021-12-01 | 2021-12-20 |

Table S3: The estimated vaccine protection rates of the partial, full and booster vaccinated against COVID-19 infection in the 6 periods considered in Figure 1.

(a) Pre-Delta period

| Country | Time | Vaccine | Partial | Full |
|---|---|---|---|---|
| Brazil | 2021-01-16 ∼ 2021-05-19 | AstraZeneca, Sinovac | 0.625 (0.04) | 0.75 ( 0.03) |
| Germany | 2020-12-26 ∼ 2021-02-28 | Janssen, Moderna, AstraZeneca, Pfizer | 0.56 (0.04) | 0.89 (0.02) |
| Italy | 2020-12-26 ∼ 2021-04-01 | Janssen, Moderna, AstraZeneca, Pfizer | 0.58 (0.04) | 0.94 (0.01) |
| Peru | 2021-02-07 ∼ 2021-06-09 | AstraZeneca, Pfizer, Sinopharm | 0.64 (0.04) | 0.76 (0.03) |
| Turkey | 2021-02-11 ∼ 2021-04-27 | Sinovac, Pfizer | 0.48 (0.04) | 0.74 (0.02) |
| UK | 2021-01-09 ∼ 2021-02-21 | AstraZeneca, Pfizer | 0.52 (0.04) | 0.68 (0.03) |
| US | 2020-12-12 ∼ 2021-02-22 | Moderna, Pfizer | 0.55 (0.04) | 0.95 (0.01) |
| Ave (SE) | | | 0.565 (0.02) | 0.816 (0.04) |

(b) Intervening I period

| Country | Time | Vaccine | Partial | Full |
|---|---|---|---|---|
| Brazil | 2021-05-20 ∼ 2021-08-15 | Pfizer, AstraZeneca, Sinovac | 0.52 (0.04) | 0.68 (0.02) |
| Germany | 2021-03-01 ∼ 2021-07-04 | Janssen, Moderna, AstraZeneca, Pfizer | 0.505 (0.04) | 0.67 (0.02) |
| Italy | 2021-04-02 ∼ 2021-07-04 | Janssen, Moderna, AstraZeneca, Pfizer | 0.55 (0.04) | 0.7 (0.03) |
| Peru | 2021-06-10 ∼ 2021-09-12 | AstraZeneca, Pfizer, Sinopharm | 0.46 (0.04) | 0.73 (0.02) |
| Turkey | 2021-04-28 ∼ 2021-06-20 | Sinovac, Pfizer | 0.235 (0.04) | 0.49 (0.03) |
| UK | 2021-02-22 ∼ 2021-05-23 | Moderna, AstraZeneca, Pfizer | 0.46 (0.04) | 0.64 (0.03) |
| US | 2021-02-23 ∼ 2021-06-20 | Janssen, Moderna, Pfizer | 0.675 (0.03) | 0.87 (0.02) |
| Ave (SE) | | | 0.486 (0.05) | 0.683 (0.04) |

(c) Delta-dominated period

| Country | Time | Vaccine | Partial | Full |
|---|---|---|---|---|
| Brazil | 2021-08-16 ∼ 2021-09-18 | Janssen, Pfizer, AstraZeneca, Sinovac | 0.385 (0.04) | 0.59 (0.02) |
| Germany | 2021-07-05 ∼ 2021-08-28 | Janssen, Moderna, AstraZeneca, Pfizer | 0.40 (0.03) | 0.60 (0.02) |
| Italy | 2021-07-05 ∼ 2021-09-18 | Janssen, Moderna, AstraZeneca, Pfizer | 0.355 (0.04) | 0.57 (0.03) |
| Peru | 2021-09-13 ∼ 2021-10-13 | AstraZeneca, Pfizer, Sinopharm | 0.415 (0.03) | 0.61 (0.02) |
| UK | 2021-05-24 ∼ 2021-09-28 | Moderna, AstraZeneca, Pfizer | 0.34 (0.03) | 0.56 (0.02) |
| US | 2021-06-21 ∼ 2021-08-11 | Janssen, Moderna, Pfizer | 0.48 (0.04) | 0.74 (0.03) |
| Ave (SE) | | | 0.396 (0.02) | 0.612 (0.03) |

Continued on next page

**Table S3 – continued from previous page**

(d) Pre-Omicron period

| Country | Time | Vaccine | Partial | Full | Booster |
|---|---|---|---|---|---|
| Brazil | 2021-09-19 ~ 2021-11-28 | Janssen, Pfizer, AstraZeneca, Sinovac | 0.28 (0.06) | 0.55 (0.02) | 0.82 (0.05) |
| Germany | 2021-08-29 ~ 2021-11-25 | Janssen, Moderna, AstraZeneca, Pfizer | 0.344 (0.07) | 0.59 (0.03) | 0.795 (0.05) |
| Italy | 2021-09-19 ~ 2021-11-25 | Janssen, Moderna, AstraZeneca, Pfizer | 0.325 (0.06) | 0.55 (0.03) | 0.82 (0.06) |
| Peru | 2021-10-14 ~ 2021-12-05 | AstraZeneca, Pfizer, Sinopharm | 0.34 (0.06) | 0.56 (0.03) | 0.824 (0.06) |
| Turkey | 2021-06-21 ~ 2021-12-05 | Sinovac, Pfizer | 0.205 (0.05) | 0.47 (0.02) | 0.788 (0.6) |
| UK | 2021-09-29 ~ 2021-11-26 | Moderna, AstraZeneca, Pfizer | 0.31 (0.06) | 0.54 (0.02) | 0.816 (0.05) |
| US | 2021-08-12 ~ 2021-11-30 | Janssen, Moderna, Pfizer | 0.46 (0.05) | 0.70 (0.03) | 0.97 (0.03) |
| Ave (SE) | | | 0.323 (0.03) | 0.566 (0.03) | 0.833 (0.02) |

(e) Intervening II period

| Country | Time | Vaccine | Partial | Full | Booster |
|---|---|---|---|---|---|
| Brazil | 2021-11-29 ~ 2022-01-02 | Janssen, Pfizer, AstraZeneca, Sinovac | 0.184 (0.04) | 0.49 (0.03) | 0.694 (0.07) |
| Germany | 2021-11-26 ~ 2022-01-02 | Janssen, Moderna, AstraZeneca, Pfizer | 0.30 (0.06) | 0.50 (0.04) | 0.70 (0.05) |
| Italy | 2021-11-26 ~ 2022-01-02 | Janssen, Moderna, AstraZeneca, Pfizer | 0.295 (0.04) | 0.53 (0.03) | 0.718 (0.05) |
| Peru | 2021-12-06 ~ 2022-01-02 | AstraZeneca, Pfizer, Sinopharm | 0.265 (0.05) | 0.51 (0.05) | 0.706 (0.06) |
| Turkey | 2021-12-06 ~ 2022-01-16 | Sinovac, Pfizer | 0.055 (0.06) | 0.37 (0.05) | 0.622 (0.06) |
| UK | 2021-11-27 ~ 2022-12-19 | Moderna, AstraZeneca, Pfizer | 0.116 (0.05) | 0.48 (0.05) | 0.688 (0.07) |
| US | 2021-12-01 ~ 2022-12-19 | Janssen, Moderna, Pfizer | 0.34 (0.04) | 0.56 (0.04) | 0.736 (0.04) |
| Ave (SE) | | | 0.222 (0.04) | 0.491 (0.02) | 0.695 (0.01) |

(f) Omicron-dominated period

| Country | Time | Vaccine | Partial | Full | Booster |
|---|---|---|---|---|---|
| Brazil | 2022-01-03 ~ 2022-03-15 | Janssen, Pfizer, AstraZeneca, Sinovac | 0.07 (0.04) | 0.38 (0.03) | 0.628 (0.06) |
| Germany | 2022-01-03 ~ 2022-03-15 | Janssen, Moderna, AstraZeneca, Pfizer | 0.115 (0.04) | 0.41 (0.05) | 0.646 (0.06) |
| Italy | 2022-01-03 ~ 2022-03-15 | Janssen, Moderna, AstraZeneca, Pfizer, Novavax | 0.16 (0.05) | 0.44 (0.04) | 0.664 (0.07) |
| Peru | 2022-01-03 ~ 2022-03-15 | AstraZeneca, Pfizer, Sinopharm | 0.10 (0.04) | 0.40 (0.03) | 0.64 (0.06) |
| Turkey | 2022-01-17 ~ 2022-03-15 | Sinovac, Pfizer, Turkovac | 0.038 (0.07) | 0.26 (0.04) | 0.556 (0.06) |
| UK | 2021-12-20 ~ 2022-03-15 | Moderna, AstraZeneca, Pfizer | 0.04 (0.06) | 0.36 (0.02) | 0.616 (0.06) |
| US | 2021-12-20 ~ 2022-03-15 | Janssen, Moderna, Pfizer | 0.285 (0.06) | 0.45 (0.04) | 0.67 (0.07) |
| Ave (SE) | | | 0.115 (0.03) | 0.386 (0.02) | 0.631 (0.01) |

Table S4: The observed and the projected numbers (in thousands), and the percentages (%) of confirmed cases (a) and deaths (b) under the two scenarios (with no and the only-partial vaccination) relative to the observed numbers during the period from the start of partial vaccination to start of booster vaccination. The 95% confidence intervals are attached to the projected numbers and percentages of the two scenarios.

(a) Confirmed cases

| Country | Observed | Scenarios | | | |
|---|---|---|---|---|---|
| | | No vaccination | | Partial vaccination | |
| | | Cases | Percentage | Cases | Percentage |
| Brazil | 12762 | 22019 (18344, 25695) | 173 (144, 201) | 14507 (13358, 15656) | 114 (105, 123) |
| Germany | 2325 | 4000 (3401, 4600) | 172 (146, 198) | 2649 (2383, 2915) | 114 (102, 125) |
| Italy | 2597 | 6980 (6413, 7546) | 269 (247, 291) | 3369 (3249, 3488) | 130 (125, 134) |
| Peru | 1006 | 1210 (1064, 1355) | 120 (106, 135) | 1070 (956, 1184) | 106 (95, 118) |
| Turkey | 2856 | 4075 (3809, 4341) | 143 (133, 152) | 3225 (3031, 3418) | 113 (106, 120) |
| UK | 4796 | 23262 (21753, 24771) | 485 (454, 516) | 14606 (13156, 16057) | 305 (274, 335) |
| US | 20341 | 98055 (88040, 108070) | 482 (433, 531) | 46954 (40654, 53253) | 231 (200, 262) |
| Total | 46683 | 159601 (142825, 176378) | 342 (306, 378) | 86380 (76788, 95972) | 185 (164, 206) |

(b) Deaths

| Country | Observed | Scenarios | | | |
|---|---|---|---|---|---|
| | | No vaccination | | Partial vaccination | |
| | | Deaths | Percentage | Deaths | Percentage |
| Brazil | 382 | 582 (494, 670) | 152 (129, 176) | 418 (385, 452) | 110 (101, 118) |
| Germany | 63 | 72 (65, 80) | 116 (104, 127) | 64 (58, 69) | 102 (93, 111) |
| Italy | 59 | 91 (87, 96) | 155 (148, 163) | 63 (61, 65) | 108 (104, 111) |
| Peru | 92 | 101 (89, 112) | 110 (98, 122) | 95 (85, 104) | 103 (93, 114) |
| Turkey | 23 | 32 (30, 34) | 140 (132, 149) | 25 (24, 27) | 112 (105, 119) |
| UK | 55 | 108 (101, 115) | 195 (182, 207) | 83 (76, 90) | 150 (137, 162) |
| US | 317 | 829 (754, 904) | 262 (238, 285) | 460 (425, 495) | 145 (134, 156) |
| Total | 990 | 1815 (1620, 2010) | 183 (164, 203) | 1208 (1114, 1302) | 122 (113, 132) |

Table S5: The observed and the projected numbers (in thousands), and the percentages (%) of confirmed cases (a) and deaths (b) under the three scenarios (with no booster, half and double the actual booster vaccination rates) relative to the observed numbers during the period from the start of booster vaccination to March 15, 2022. The 95% confidence intervals are attached to the projected numbers and percentages of the three scenarios.

(a) Confirmed cases

| Country | Observed | Scenarios | | | | | |
|---|---|---|---|---|---|---|---|
| | | No booster | | Half booster | | Double booster | |
| | | Cases | Percentage | Cases | Percentage | Cases | Percentage |
| Brazil | 8239 | 11598 (10207, 12989) | 141 (124, 158) | 9887 (8849, 10926) | 120 (107, 133) | 6528 (5751, 7306) | 79 (70, 89) |
| Germany | 13668 | 18589 (16773, 20404) | 136 (123, 149) | 15369 (14274, 16463) | 112 (104, 120) | 8322 (7688, 8956) | 61 (56, 66) |
| Italy | 8913 | 12452 (11990, 12914) | 140 (135, 145) | 10124 (9765, 10483) | 114 (110, 118) | 5583 (5014, 6152) | 63 (56, 69) |
| Peru | 1349 | 1798 (1398, 2197) | 133 (104, 163) | 1591 (1317, 1865) | 118 (98, 138) | 1136 (950, 1322) | 84 (70, 98) |
| Turkey | 9160 | 14961 (12312, 17610) | 163 (134, 192) | 11857 (10639, 13075) | 129 (116, 143) | 5852 (4617, 7086) | 64 (50, 77) |
| UK | 12113 | 15692 (14779, 16605) | 130 (122, 137) | 12588 (12077, 13098) | 104 (100, 108) | 5851 (5266, 6436) | 48 (43, 53) |
| US | 43056 | 56268 (52582, 59955) | 131 (122, 139) | 49670 (47601, 51738) | 115 (111, 120) | 35548 (33089, 38006) | 83 (77, 88) |
| Total | 96498 | 131358 (120041, 142675) | 136 (124, 148) | 111085 (104522, 117648) | 115 (108, 122) | 68820 (62374, 75265) | 71 (65, 78) |

(b) Deaths

| Country | Observed | Scenarios | | | | | |
|---|---|---|---|---|---|---|---|
| | | No booster | | Half booster | | Double booster | |
| | | Deaths | Percentage | Deaths | Percentage | Deaths | Percentage |
| Brazil | 65 | 82 (73, 92) | 127 (113, 142) | 74 (66, 82) | 114 (102, 126) | 57 (51, 62) | 87 (78, 96) |
| Germany | 34 | 40 (35, 45) | 120 (105, 135) | 36 (33, 39) | 105 (96, 115) | 27 (25, 29) | 79 (73, 85) |
| Italy | 27 | 36 (34, 38) | 134 (125, 142) | 30 (28, 31) | 111 (105, 116) | 18 (16, 20) | 66 (58, 74) |
| Peru | 12 | 15 (12, 18) | 127 (102, 153) | 14 (11, 16) | 115 (95, 135) | 10 (9, 12) | 87 (75, 99) |
| Turkey | 47 | 72 (60, 84) | 153 (128, 178) | 59 (53, 64) | 125 (113, 136) | 33 (27, 38) | 70 (58, 81) |
| UK | 27 | 33 (31, 35) | 124 (116, 132) | 27 (25, 29) | 101 (95, 107) | 15 (14, 16) | 55 (51, 60) |
| US | 350 | 425 (404, 447) | 122 (115, 128) | 388 (373, 404) | 111 (106, 115) | 308 (294, 322) | 88 (84, 92) |
| Total | 561 | 704 (649, 759) | 126 (116, 135) | 627 (590, 664) | 112 (105, 118) | 467 (435, 499) | 83 (78, 89) |

Table S6: The peak (maximum) values of the observed and the projected numbers (in millions) of active confirmed cases during the post-vaccine period with the days that the projected numbers of active confirmed cases under each scenario would exceed the observed peak. The percentages (%) of the projected peak values over the observed peak values are provided in parentheses. The five scenarios are (i) no vaccination at all; (ii) only partial vaccination; (iii) no booster, (iv) half and (v) twice the booster up-take.

| Country | Observed | Scenarios | | | | | | | | | |
|---|---|---|---|---|---|---|---|---|---|---|---|
| | | No vaccination | | Partial vaccination | | No booster | | Half booster | | Double booster | |
| | Peak | Peak | Days | Peak | Days | Peak | Days | Peak | Days | Peak | Days |
| Brazil | 1.89 | 11.82 (624) | 76 | 12.07 (637) | 71 | 2.62 (138) | 36 | 2.19 (116) | 24 | 1.31 (69) | 0 |
| Germany | 2.53 | 6.35 (251) | 71 | 6.27 (248) | 65 | 3.53 (140) | 41 | 2.94 (116) | 27 | 1.38 (54) | 0 |
| Italy | 1.97 | 3.44 (175) | 70 | 5.83 (296) | 66 | 2.86 (145) | 36 | 2.23 (113) | 21 | 1.1 (56) | 0 |
| Peru | 0.41 | 3.85 (940) | 79 | 2.86 (697) | 71 | 0.51 (123) | 23 | 0.44 (108) | 14 | 0.3 (74) | 0 |
| Turkey | 1.20 | 2.77 (231) | 111 | 2.11 (176) | 156 | 2.12 (178) | 59 | 1.6 (134) | 42 | 0.66 (55) | 0 |
| UK | 1.76 | 4.04 (230) | 80 | 2.13 (121) | 76 | 2.43 (138) | 51 | 1.74 (99) | 0 | 0.65 (37) | 0 |
| US | 8.29 | 31.89 (385) | 76 | 20.94 (253) | 72 | 11.2 (135) | 31 | 9.45 (114) | 21 | 5.68 (69) | 0 |

Table S7: The estimated vaccine protection rates of the partial, full and booster vaccinated against COVID-19 infection in the intervening II and Omicron-dominated periods in the sensitivity analyses of the symptomatic rate and the average time duration from recovery to loss of natural immunity. Compared to the results in Table S3 (e) and (f), the largest difference between VPRs in the main analysis and those in sensitivity analysis of asymptomatic rate was 8.6% for the partial vaccination in the Omicron-dominated period in Italy, for other vaccinations, periods and countries the differences were no more than 3.4%, and the average of the absolute differences was 1.26% (SE: 0.23%). And the largest difference between VPRs in the main analysis and those in the sensitivity analysis of duration of natural immunity was 5% for the full vaccination in the Intervening II period in Turkey, for other vaccinations, periods and countries the differences were no more than 3%, and the average of the absolute differences was 0.94% (SE: 0.17%).

| Country | Asymptomatic rate | | | | | | Time from recovery to loss of natural immunity | | | | | |
|---|---|---|---|---|---|---|---|---|---|---|---|---|
| | Intervening II | | | Omicron-dominated | | | Intervening II | | | Omicron-dominated | | |
| | Partial | Full | Booster | Partial | Full | Booster | Partial | Full | Booster | Partial | Full | Booster |
| Brazil | 0.167 | 0.51 | 0.706 | 0.07 | 0.38 | 0.628 | 0.20 | 0.50 | 0.70 | 0.085 | 0.39 | 0.634 |
| Germany | 0.28 | 0.52 | 0.712 | 0.115 | 0.41 | 0.646 | 0.28 | 0.52 | 0.712 | 0.10 | 0.40 | 0.64 |
| Italy | 0.28 | 0.52 | 0.712 | 0.246 | 0.42 | 0.652 | 0.295 | 0.53 | 0.718 | 0.13 | 0.42 | 0.652 |
| Peru | 0.235 | 0.49 | 0.694 | 0.10 | 0.40 | 0.64 | 0.265 | 0.51 | 0.706 | 0.07 | 0.38 | 0.628 |
| Turkey | 0.085 | 0.39 | 0.634 | 0.051 | 0.27 | 0.562 | 0.072 | 0.42 | 0.594 | 0.038 | 0.26 | 0.556 |
| UK | 0.15 | 0.50 | 0.70 | 0.055 | 0.37 | 0.622 | 0.116 | 0.48 | 0.688 | 0.04 | 0.36 | 0.616 |
| US | 0.34 | 0.56 | 0.736 | 0.298 | 0.46 | 0.676 | 0.34 | 0.56 | 0.736 | 0.298 | 0.46 | 0.676 |
| Average | 0.220 | 0.499 | 0.699 | 0.134 | 0.387 | 0.632 | 0.224 | 0.503 | 0.693 | 0.109 | 0.381 | 0.629 |

### S7 Figures

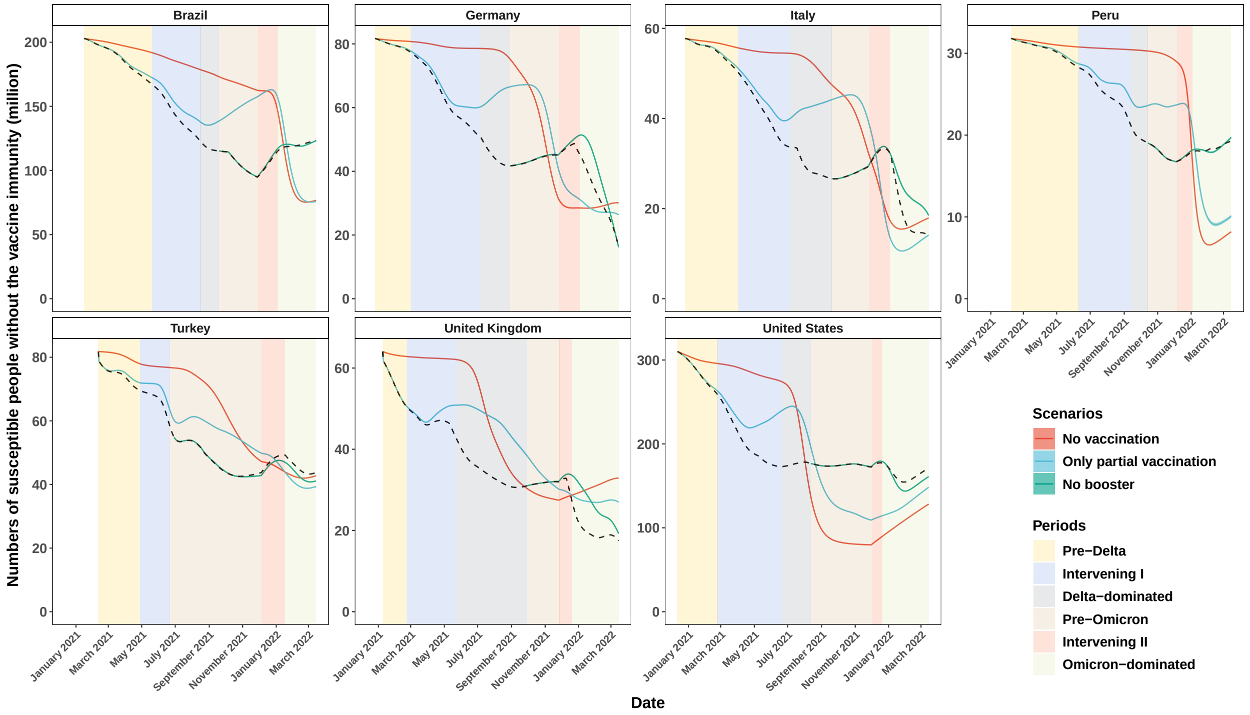


Figure S1: The projected daily numbers (in millions) of susceptible people with no vaccine immunity under the three vaccination scenarios (color curves) and the imputed ones using real data (black dashed lines). The 95% confidence bands of the projected numbers are indicated by colored areas.

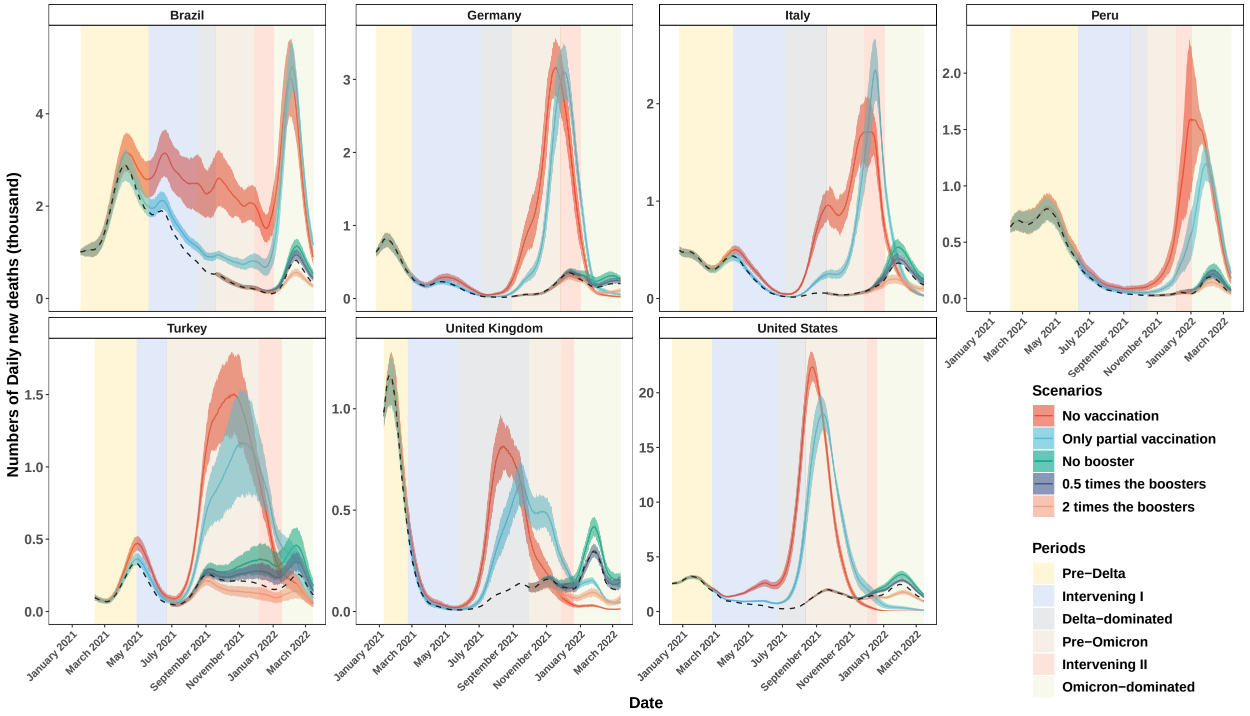


Figure S2: The actual (black dashed lines), and the projected numbers (in thousands) of daily new deaths (color curves) and their 95% confidence bands (color area) under the five vaccination scenarios.

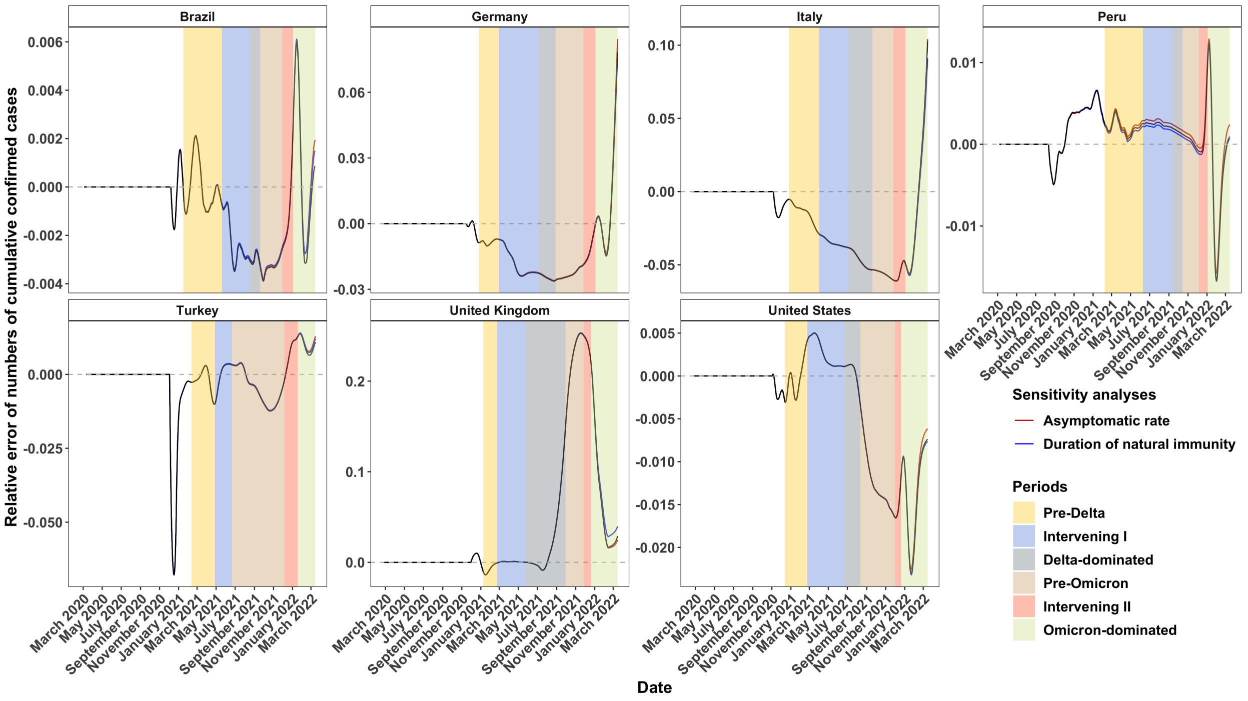


Figure S3: The relative errors between the mean projected numbers of cumulative confirmed cases by 1000 simulations with the estimated parameters and the observed numbers of cumulative confirmed cases for the main analysis (black), and sensitivity analyses for the asymptomatic rate (red) and the duration of natural immunity (blue) in the seven countries.

18. Tsybakov, A. *Introduction to Nonparametric Estimation* (Springer, 2009).
